# Flow-gradient Phenotypes and Functional Recovery After Transcatheter Aortic Valve Implantation for Severe Aortic Stenosis: A COMPARE-TAVI 1 Sub-study

**DOI:** 10.64898/2026.05.04.26352421

**Authors:** Emil Johannes Ravn, Henrik Vase, Henrik Nissen, Kristian Hejlesen, Karen Juel Andersen, Nils Sofus Borg Mogensen, Rasmus Carter-Storch, Anne Dybro, Troels Thim, Philip Freeman, Frederik Uttenthal, Ulrik Christiansen, Evald Høj Christiansen, Christian Juhl Terkelsen, Jordi Sanchez Dahl

## Abstract

**BACKGROUND:** Patients with severe aortic stenosis (AS) exhibit heterogeneous flow-gradient hemodynamics and ventricular remodeling, which may influence symptomatic, functional, and structural responses to transcatheter aortic valve implantation (TAVI). Thus, we evaluated differences in functional recovery and reverse remodeling after transfemoral TAVI across flow-gradient phenotypes.

**METHODS:** In this sub-study of the COMPARE-TAVI 1 trial, 975 patients undergoing transfemoral TAVI were classified as classical low-flow low-gradient (cLFLG, 9.1%), paradoxical low-flow low-gradient (pLFLG, 7.7%), low-flow high-gradient (24.7%), normal-flow low-gradient (NFLG, 13.0%), and normal-flow high-gradient (45.4%). The primary functional outcome was longitudinal change in six-minute walk test distance (6MWTD) from baseline to 1 year follow-up. Secondary endpoints included changes in NYHA functional class and reverse remodeling from baseline to 1 year follow-up along with the incidence and risk of all-cause death and a composite MACE-endpoint.

**RESULTS:** Mean 6MWTD increased by 59±4 meters at 1-month (p=0.000) with no additional improvement at 1-year, but with heterogeneity between groups (p=0.000). Improvements among NFLG, cLFLG and low-flow high-gradient AS were comparable with normal-flow high-gradient AS, while pLFLG AS exhibited significantly increments at 1-year (−28±15 meters, p=0.007). Patients with NFLG, cLFLG and pLFLG were more symptomatic at baseline (NYHA ≥III: 40.5%, 57.3% and 50.6%, respectively, p=0.000). NYHA improved in all groups at 1-year follow-up (p=0.000), although persistent symptoms at 1-year were most frequent in pLFLG (NYHA ≥II; 38.7%, p=0.012). Reverse remodeling was also comparable between normal-flow high-gradient AS and NFLG, cLFLG, and low-flow high-gradient AS, respectively, but attenuated in pLFLG AS in both unadjusted and adjusted analyses. No differences were observed in the incidence and risk of all-cause death or the composite MACE-endpoint.

**CONCLUSION:** TAVI associates with functional recovery across all flow-gradient phenotypes, although with heterogeneous responses. Patients with NFLG showed comparable functional recovery and reverse remodeling at 1-year follow-up compared with normal-flow high-gradient AS, whereas pLFLG demonstrated attenuated benefits across all parameters.

## INTRODUCTION

Aortic stenosis (AS) is characterized by left ventricular (LV) pressure overload leading to LV hypertrophy, concentric remodeling, and reduced LV systolic function. Although severe AS typically has been defined as aortic valve area (AVA) <1.0 cm^2^ with transvalvular mean gradient ≥40 mmHg, there has been an increasing recognition that AS may be considered severe despite low gradients.^1^ This view is largely driven by cohort-studies demonstrating that patients with low-gradient AS defined as mean-gradient <40 mmHg and AVA<1.0 cm^2^ is associated with a poor prognosis if treated conservatively, in particular in the presence of a reduced flow (stroke volume index (SVi) <35 mL/m^2^).^2,3^

While a low-flow condition may be the consequence of a reduced LV ejection fraction (LVEF), it may also reflect a distinct remodeling process, ensuing a small, restrictive LV cavity with reduced LV systolic longitudinal function,^4^ unable to preserve flow despite preserved LVEF.^2,5^ However, numerous studies have demonstrated that reduced SVi may preclude symptom improvement after aortic valve replacement, but few have evaluated the impact of flow-gradient patterns on functional recovery following transcatheter aortic valve implantation (TAVI),^6^ leading to a paucity in data delineating the possible mechanism. Furthermore, a substantial number of patients referred for aortic valve replacement present with low gradients and AVA<1.0 cm^2^ despite preserved SVi ≥35 mL/m^2^ (normal-flow low-gradient (NFLG) AS).^7^ While some have regarded this condition to represent a severe AS phenotype with low gradients attributed to the coexistence of reduced aortic compliance and systemic hypertension,^8^ NFLG patients share similar survival rates with moderate AS.^9^ Accordingly, despite studies suggesting that these patients may benefit from aortic valve replacement, guidelines consider this entity to represent moderate AS.^7,10,11^ Thus, the aim of this study was to investigate the impact of different flow-gradient patterns on functional recovery, reverse LV remodeling, and adverse outcomes following transfemoral TAVI.

## METHODS

The COMPARE-TAVI 1 trial is a multi-center, prospective, randomized controlled trial, in which all-comer patients were randomized 1:1 to receive transfemoral TAVI with either Sapien 3/Sapien 3 Ultra Transcatheter Heart Valve (THV) series (Edwards Lifesciences, Irvine, California, USA) or the Myval/Myval Octacor THV series (Meril Life Sciences Pvt. Ltd., Vapi, Gujarat, India).^12^ The study was approved by the Ethics Committee of the Central Denmark Region. Written and verbal informed consent was obtained from all screened participants at the time of enrollment. The study was conducted in accordance with the Declaration of Helsinki, and the study protocol has previously been published.^12^

Patients underwent a clinical examination with assessment of six-minute walk test distance (6MWTD) and New York Heart Association (NYHA) functional class, including an echocardiogram prior to TAVI (baseline), at 1-month and 1-year follow-up. In the present sub-study, we excluded patients with missing SVi (n=7), LVEF (n=2), AVA (n=3), and aortic mean-gradient (n=1) at baseline along with patients, who had concomitant moderate-severe aortic regurgitation (n=3) or previous aortic valve replacement (n=40).

### Echocardiography

A comprehensive transthoracic echocardiography was performed according to a prespecified core laboratory echocardiographic protocol using Vivid 9 (GE Healthcare, Horten, Norway) or EPIQ7 (Philips Professional Healthcare, Netherlands) machines. Examinations were digitally archived locally, de-identified and transferred in raw format to a core laboratory. Echocardiograms were assigned to 5 blinded readers that included highly experienced research fellows and board-certified cardiologists with level III certification in echocardiography and were subsequently approved by the core-lab director. Furthermore, interobserver analysis within the core laboratory demonstrated acceptable variability between analysts, with a variance of ±8–15% key parameters such as SVi and LVEF. Echocardiograms were analyzed using Viewpoint 6 (GE Healthcare, Horten, Norway) and Intellispace Cardiovascular (Philips Healthcare Best, Netherlands) software. Echocardiographic measurements and definitions have already been described in our previously published sub-study on the COMPARE-TAVI 1 cohort.^13^

In this sub-study, patients were stratified into five groups according to flow-gradient pattern. Low-flow low-gradient (AVA <1.0 cm^2^, SVi <35 mL/m^2^, and mean-gradient <40 mmHg) was further subdivided according to LVEF in classical low-flow low-gradient (cLFLG, LVEF <50%) or paradoxical low-flow low-gradient (pLFLG, LVEF ≥50%) AS; NFLG (AVA <1.0 cm^2^, SVi >35 mL/m^2^, and mean-gradient <40 mmHg); low-flow high-gradient (AVA <1.0 cm^2^, SVi <35 mL/m^2^, and mean-gradient >40 mmHg), and normal-flow high-gradient (AVA <1.0 cm^2^, SVi >35 mL/m^2^, and mean-gradient >40 mmHg) AS.

The primary outcome was defined as improvement in 6MWTD across flow-gradient phenotypes from baseline to 1-year follow-up. Secondary outcomes included changes in NYHA functional class and structural reverse remodeling, evaluated by changes in LV mass index, LV end-diastolic diameter, relative wall thickness, and LVEF from baseline to 1-year follow-up, as well as the 1-year risk of all-cause death and a composite endpoint comprising all-cause death, stroke, ≥moderate aortic regurgitation, and ≥moderate hemodynamic valve deterioration. No patients were lost to follow-up.

### Statistics

Normally distributed continuous variables are presented as mean ± standard deviation, and categorical variables as counts with percentages. Normality of continuous variables was assessed using quantile–quantile plots and the Shapiro–Wilk test. Comparisons of baseline characteristics across flow–gradient phenotypes were performed using one-way analysis of variance for continuous variables and Pearson’s chi-square test for categorical variables. Comparisons of echocardiographic characteristics were performed using one-way analysis of variance.

The distribution of NYHA functional class across flow-gradient phenotypes was compared cross-sectionally using chi-square tests at baseline, 1-month follow-up, and 1-year follow-up. Additional dichotomized analyses according to NYHA functional class ≥III and ≥II were performed between groups.

Longitudinal changes in 6MWTD and reverse remodeling parameters were analyzed using mixed-effects linear regression models, with time, flow-gradient phenotype, and their interaction specified as fixed effects. Normal-flow high-gradient AS served as the reference group. A random intercept was included to account for repeated measurements within individuals, and model parameters were estimated using maximum likelihood estimation. Model fit was assessed using the log-likelihood. Adjusted marginal predictions were generated to visualize phenotype-specific trajectories at each time point. Analyses of 6MWTD were additionally adjusted for clinically relevant baseline characteristics according to significant differences between groups.

Three additional sensitivity analyses were performed for 6MWTD, excluding patients with incomplete follow-up, patients with atrial fibrillation, and patients with ischemic heart disease. Time-to-death was calculated as the time from the date of the TAVI procedure to the date of death from all-causes. Time-to-event analyses were performed using non-parametric Kaplan-Meier methods with log-rank tests, as well as semi-parametric Cox proportional hazards regression. The proportional hazards assumption was assessed using Schoenfeld residuals.

All statistical analysis were conducted using STATA (StataCorp. 2024. Stata Statistical Software: Version 18.0. College Station, Texas, USA).

## RESULTS

### Baseline characteristics

From June 2020 to November 2023, 1,031 patients were included in the COMPARE-TAVI 1 trial. According to the exclusion criteria of this sub-study, 56 participants were subsequently excluded, leaving a total of 975 participants. Patients were grouped according to flow-gradient phenotype; cLFLG (n=89, 9%), pLFLG (n=75, 8%), low-flow high-gradient (n=241, 25%), NFLG (n=127, 13%), and normal-flow high-gradient (n=443, 45%) (Figure 1).

**Figure 1.**
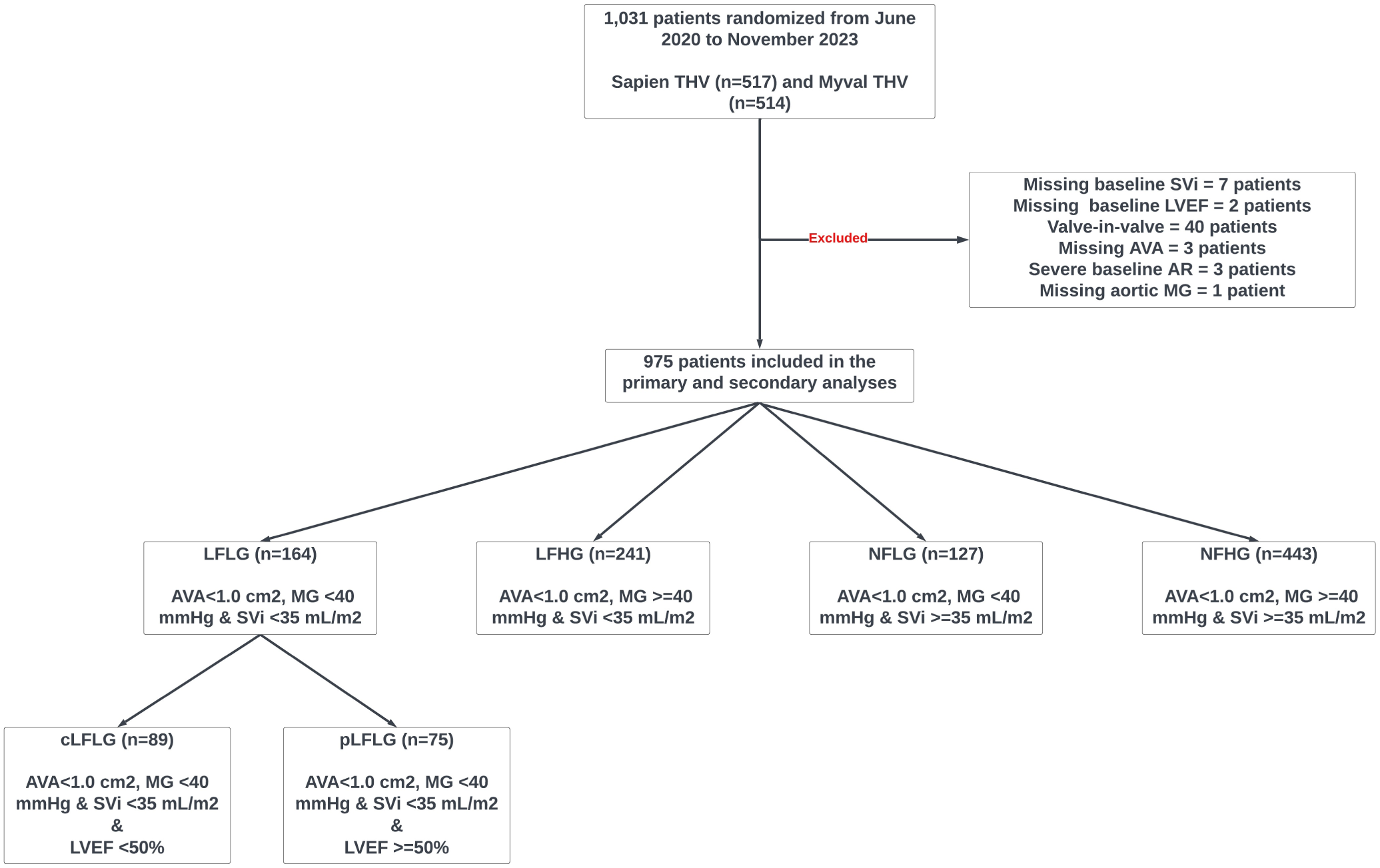
– Flowchart describing the population included in this study and stratification according to flow-gradient phenotypes. Abbreviations: AR, aortic regurgitation; AVA, aortic valve area; cLFLG, classical low flow-low gradient; LFHG, low flow-high gradient; LFLG, low flow-low gradient; LVEF, left ventricular ejection fraction; MG, mean gradient; NFLG, normal flow-low gradient; NFHG, normal flow-high gradient; pLFLG, paradoxical low flow-low gradient; SVi, stroke volume index; THV, transcatheter heart valve.

Patients with cLFLG and low flow-high gradient AS had significantly lower pre-TAVI higher heart rate (cLFLG 82 (SD 18) vs. pLFLG 75 (SD 14) vs. LFHG 79 (SD 15) vs. NFLG 69 (SD 12) vs. normal-flow high-gradient 71 (SD 12); p=0.000), while patients with cLFLG AS also had lower systolic blood pressure (cLFLG 125 (SD 21) mmHg vs. pLFLG 138 (SD 19) mmHg vs. LFHG 136 (SD 19) mmHg vs. NFLG 142 (SD 19) mmHg vs. normal-flow high-gradient 143 (SD 21) mmHg; p=0.000). A medical history of previous myocardial infarction (p=0.012), diabetes (p=0.017), pacemaker (p=0.011), and congestive heart failure (p=0.000) was most frequent among patients with cLFLG, while atrial fibrillation (p=0.000) and NYHA class ≥III (p=0.000) was more frequent among patients with cLFLG and pLFLG AS, with no difference between the two groups (Table 1).

**Table 1.** – baseline characteristics.

| Variables | cLFLG | pLFLG | LFHG | NFLG | NFHG | p-value |
| --- | --- | --- | --- | --- | --- | --- |
| N | 89 (9.1) | 75 (7.7) | 241 (24.7) | 127 (13.0) | 443 (45.4) | **0.000** |
| Male sex | 65 (73.0)† | 36 (48.0) | 150 (62.2) | 73 (57.5) | 250 (56.4) | **0.010** |
| Age, years | 80 (7) | 82 (6) | 81 (7) | 81 (5) | 81 (6) | 0.49 |
| Body mass index, kg/m<sup>2</sup> | 26.8 (4.4) | 27.9 (4.6) | 28.0 (6.0) | 27.9 (4.6) | 27.1 (4.7) | 0.07 |
| Body surface area, m<sup>2</sup> | 1.93 (0.22)† | 1.90 (0.21) | 1.93 (0.24) | 1.90 (0.18) | 1.87 (0.21) | **0.004** |
| Systolic BP, mmHg | 125 (21)† | 138 (19) | 136 (19) | 142 (21) | 143 (21) | **0.000** |
| Diastolic BP, mmHg | 74 (12) | 77 (11) | 75 (12) | 74 (13) | 74 (12) | 0.30 |
| Heart-rate | 82 (18)† | 75 (14) | 79 (15) | 69 (12) | 71 (12) | **0.000** |
| Hypertension | 63 (70.8) | 60 (80.0) | 178 (73.9) | 105 (82.7)* | 327 (74.0) | 0.18 |
| Diabetes mellitus | 30 (33.7)† | 18 (24.0) | 56 (23.2) | 28 (22.0) | 79 (17.8) | **0.017** |
| Previous myocardial infarction | 15 (16.9)† | 10 (13.3) | 19 (7.9) | 20 (15.7)* | 36 (8.1) | **0.012** |
| Atrial fibrillation | 52 (58.4) | 42 (56.0)‡ | 103 (42.7) | 44 (34.6)* | 107 (24.2) | **0.000** |
| Congestive heart failure | 60 (67.4)† | 16 (21.3)‡ | 52 (21.6) | 21 (16.5)* | 40 (9.0) | **0.000** |
| NYHA class ≥3 | 51 (57.3)† | 38 (50.6)‡ | 108 (44.8) | 52 (40.9)* | 135 (30.5) | **0.000** |
| Peripheral vascular disease | 8 (9.0)† | 6 (8.0) | 17 (7.1) | 13 (10.3)* | 21 (4.8) | 0.18 |
| Previous stroke | 9 (10.1) | 8 (10.7) | 35 (14.5) | 25 (19.7) | 74 (16.7) | 0.22 |
| COPD | 13 (14.8) | 12 (16.2) | 44 (18.3) | 19 (15.1) | 65 (14.8) | 0.81 |
| Pacemaker | 19 (21.3)† | 10 (13.3) | 19 (7.9) | 17 (13.4) | 35 (7.9) | **0.011** |

\* p<0.05 for pairwise comparison: NFLG vs. NFHG
† p<0.05 for pairwise comparison: cLFLG vs. NFHG
‡ p<0.05 for pairwise comparison: pLFLG vs. NFHG
Abbreviations: BP, blood pressure; BSA, body surface area; cLFLG, classical low flow-low gradient; COPD, chronic
obstructive pulmonary disease; LFHG, low flow-high gradient; NFLG, normal flow-low gradient; NFHG, normal flow-
high gradient; pLFLG, paradoxical low flow-low gradient; Significant differences (p<0.05) are highlighted in **bold**.

While all patients had severe AS, AVA was significantly higher among patients with NFLG AS (cLFLG 0.66±0.15 cm2 vs. pLFLG 0.64±0.14 cm2 vs. low-flow high-gradient 0.54±0.12 cm2 vs. NFLG 0.93±0.18 cm2 vs. normal-flow high-gradient 0.76±0.18 cm2; p=0.000) (Table 2). Patients with cLFLG AS had the highest LV mass index and the lowest relative wall thickness, and consequently the highest prevalence of eccentric hypertrophy (cLFLG 20.2% vs. pLFLG 12.0% vs. low-flow high-gradient 12.0% vs. NFLG 11.8% vs. normal-flow high-gradient 8.1%, p=0.004) (Table 2). There was no difference in left atrial volume index (p=0.170), but patients with low-flow low-gradient AS showed lower tricuspid annular plane systolic excursion (cLFLG 19 (SD 5) mm vs. pLFLG 20 (SD 5) mm vs. low-flow high-gradient 21 (SD 5) mm vs. NFLG 23 (SD 5) mm vs. normal-flow high-gradient 24 (SD 5) mm, p=0.000), while patients with cLFLG AS had higher tricuspid regurgitation gradients (p=0.004) and pulmonary artery systolic pressures (p=0.000).

**Table 2.** – echocardiographic characteristics at baseline.

| Variables | cLFLG | pLFLG | LFHG | NFLG | NFHG | p-value |
| --- | --- | --- | --- | --- | --- | --- |
| N | 89 (9.1) | 75 (7.7) | 241 (24.7) | 127 (13.0) | 443 (45.4) | **0.000** |
| Aortic peak velocity, cm/sec | 341 (38.6)† | 358 (33.2)‡ | 445 (42.3) | 369 (35.2)* | 460 (50.7) | **0.000** |
| Aortic mean gradient, mmHg | 30.0 (6.1)† | 32.5 (5.4)‡ | 52.8 (10.0) | 33.5 (5.5)* | 56.8 (19.5) | **0.000** |
| AVA, cm<sup>2</sup> | 0.66 (0.15)† | 0.64 (0.14)‡ | 0.54 (0.12) | 0.93 (0.18)* | 0.76 (0.18) | **0.000** |
| LV ejection fraction, % | 35.7 (10.0)† | 59.3 (11.3) | 53.2 (13.7) | 56.6 (13.3)* | 59.6 (10.7) | **0.000** |
| Global longitudinal strain, % | 8.7 (2.8)† | 12.7 (2.7)‡ | 11.5 (3.4) | 13.6 (3.6) | 14.3 (3.0) | **0.000** |
| LV end-diastolic diameter, mm | 52 (7.1)† | 45 (7.4) | 46 (7.2) | 47 (6.8)* | 46 (7.1) | **0.000** |
| LV end-systolic diameter, mm | 44 (8.6)† | 33 (8.7) | 35.5 (8.5) | 35 (9.7)* | 33 (8.9) | **0.000** |
| Stroke volume index, mL/m<sup>2</sup> | 25.9 (6.1)† | 28.0 (5.6)‡ | 29.4 (4.4) | 44 (8.7)* | 46.3 (8.9) | **0.000** |
| LV mass index, g/m<sup>2</sup> | 133 (29.4)† | 110 (21.9)‡ | 122 (30.9) | 117 (32.4) | 121 (31.1) | **0.000** |
| Relative wall thickness, % | 49 (14)† | 55 (16) | 57 (19) | 51 (15)* | 57 (18) | **0.000** |
| Remodeling type, n (%) |  |  |  |  |  | **0.004** |
| - Normal geometry | 10 (11.2) | 9 (12.0) | 14 (5.8) | 16 (12.6) | 40 (9.0) |  |
| - Concentric remodeling | 8 (9.0) | 23 (30.7) | 62 (25.7) | 31 (24.4) | 112 (25.3) |  |
| - Eccentric hypertrophy | 18 (20.2) | 9 (12.0) | 29 (12.0) | 15 (11.8) | 36 (8.1) |  |
| - Concentric hypertrophy | 53 (59.6) | 34 (45.3) | 136 (56.4) | 65 (51.2) | 255 (57.6) |  |
| LA volume index (mL/m<sup>2</sup>) | 48 (18.3) | 43 (18.3) | 46 (19.4) | 43 (18.3) | 46 (16.6) | 0.17 |
| E velocity, cm/sec | 95 (32.3) | 87 (31.5) | 96 (30.8) | 90 (28.2) | 94 (31.2) | 0.23 |
| A velocity, cm/sec | 81 (42.5)† | 84 (44.9)‡ | 92 (41.3) | 98 (32.0) | 102 (33.6) | **0.000** |
| Deceleration time, ms | 179 (64.3)† | 234 (98.5) | 226 (126) | 258 (98.3) | 256 (105) | **0.000** |
| E/e lateral | 16.8 (9.3) | 14.2 (9.9) | 17.3 (11.1) | 15 (8.7) | 15.6 (7.6) | 0.12 |
| TR gradient, mmHg | 31 (15.3)† | 28 (11.6) | 32 (13.4) | 27 (11.8) | 28 (12.4) | **0.004** |
| TAPSE, mm | 19 (5)† | 20 (5)‡ | 21 (5) | 23 (5) | 24 (5) | **0.000** |
| PASP, mmHg | 43 (16.9)† | 36 (12.9) | 37 (14.5) | 34 (13.7) | 34 (13.7) | **0.000** |

\* p<0.05 for pairwise comparison: NFLG vs. NFHG
† p<0.05 for pairwise comparison: cLFLG vs. NFHG
‡ p<0.05 for pairwise comparison: pLFLG vs. NFHG
Abbreviations: cLFLG, classical low flow-low gradient; LA, left atrial; LFHG, low flow-high gradient; LV, left ventricular; NFLG, normal flow-low gradient; NFHG, normal flow-high gradient; pLFLG, paradoxical low flow-low gradient; PASP, pulmonary artery systolic pressure; TAPSE, tricuspid annular plane systolic excursion; TR, tricuspid regurgitation; Significant differences (p<0.05) are highlighted in **bold**.

### Outcomes

During a total 1-year follow-up time after TAVI, 59 patients died (cLFLG (n=7, 7.9%), pLFLG (n=4, 5.3 %), low-flow high-gradient (n=19, 7.9%), NFLG (n=7, 5.5%), and normal-flow high-gradient (n=20, 4.5%)) with no difference in rates of 1-year all-cause death or risk of all-cause death between groups (log-rank p=0.403) (Figure 2). There was also no difference between groups for the composite endpoint of all-cause death, stroke, ≥moderate aortic regurgitation, and ≥moderate hemodynamic valve deterioration (cLFLG (n=10, 11.2%), pLFLG (n=8, 10.7%), low-flow high gradient (n=41, 17.0%), NFLG (n=18, 14.2%), and normal-flow high-gradient (n=53, 12.0%; p=0.352).

**Figure 2.**
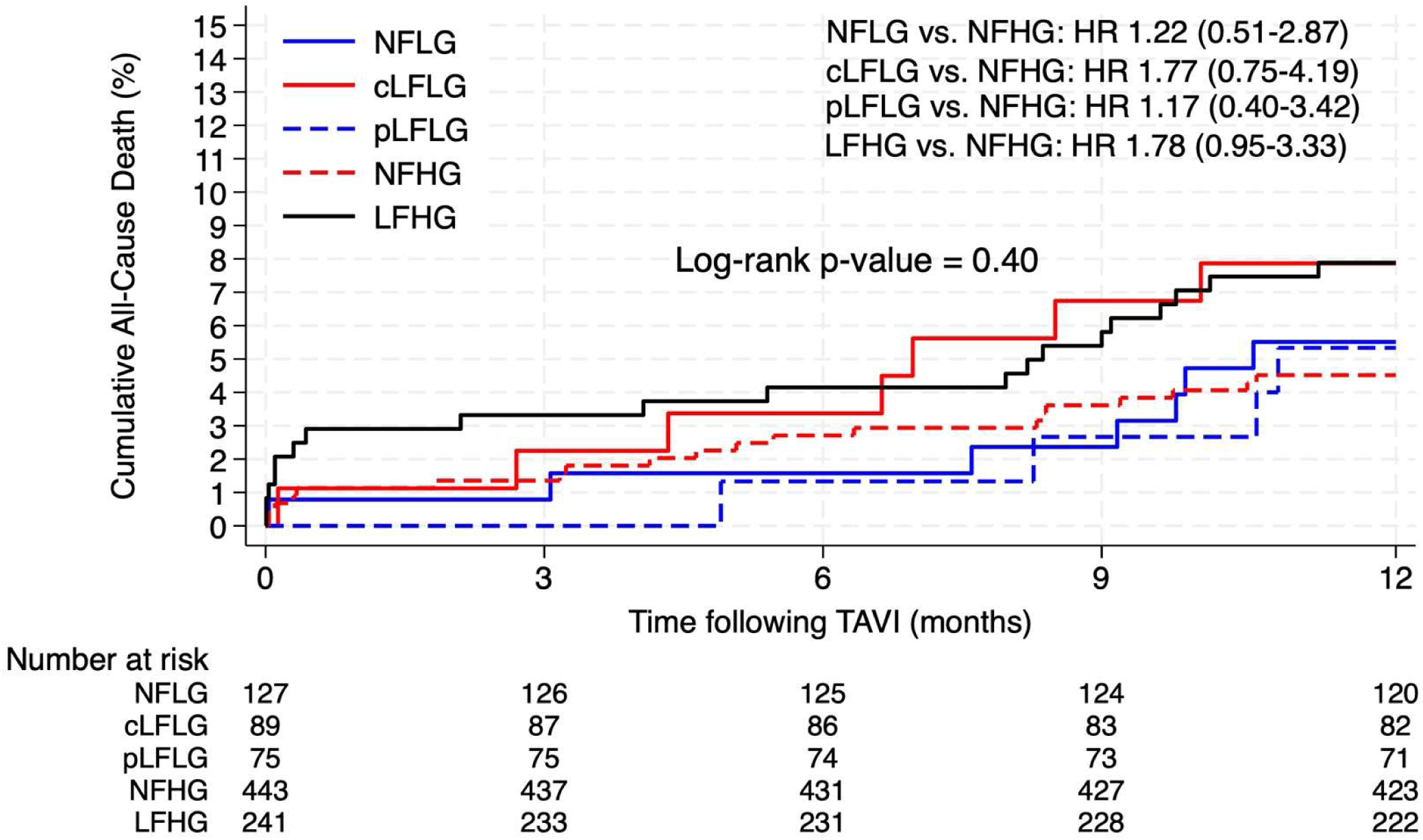
– Kaplan Meier plot illustrating risk of all-cause death at 1-year follow-up. Abbreviations: cLFLG, classical low flow-low gradient; LFHG, HR, Hazard Ratio; low flow-high gradient; NFLG, normal flow-low gradient; NFHG, normal flow-high gradient; pLFLG, paradoxical low flow-low gradient; TAVI, transcatheter aortic valve implantation.

Baseline 6MWTD was available in 648 patients (66.4%), and in 549 (56.3%) and 503 (51.6%) patients at 1-month and 1-year follow up, respectively. Overall, baseline 6MWTD was 315±4 meters, with significant differences between groups (cLFLG 294±15 vs. pLFLG 306±15 vs. low-flow high-gradient 301±9 vs. NFLG 311±11 vs. normal-flow high-gradient 329±6 meters, p=0.000). During follow-up, 6MWTD increased to 369±4 meters at 1-month and to 370±4 meters at 1-year follow up (p=0.000) (Central illustration). Improvements occurred uniformly at 1-month follow-up across all groups (59±4 meters, p=0.000), with no further improvement after 1 year (59±4 meters vs. 61±5 meters, p=0.770), but with significant heterogeneity between groups (p=0.000). Improvements in 6MWTD among NFLG AS was comparable to those observed in patients with normal-flow high-gradient AS at both 1-month (47±11 vs. 59±6 meters, p=0.180) and 1 year (54±12 vs. 61±6 meters, p=0.710), with similar improvements observed in patients with cLFLG and low-flow high-gradient AS. However, improvement in 6MWTD was less prominent in pLFLG AS compared to normal-flow high-gradient AS 1-year after TAVI (28±15 vs. 61±6 meters, p=0.007). Combined with differences in 6MWTD at baseline and the heterogenic response to TAVI, pLFLG demonstrated the lowest 6MWTD at 1-year follow up (cLFLG 350±15 vs pLFLG 334±15 vs. low-flow high-gradient 351±9 vs. NFLG 365±12 vs. normal-flow high-gradient 390±6 meters, p=0.000).

After multivariable adjustment for clinically relevant baseline characteristics (e.g., sex, age, BSA, diabetes mellitus, prior myocardial infarction, atrial fibrillation, congestive heart failure, NYHA class, and pacemaker), differences in baseline 6MWTD between groups were no longer significant. Patients with NFLG still shared similar improvements in 6MWTD with normal-flow high-gradient AS, while pLFLG AS was still associated with attenuated improvements following TAVI, with significantly smaller increments in 6MWTD at 1 month (–29 m, p=0.014) and 1 year (– 32 m, p=0.009) (Figure 3). We also performed a multivariable-adjusted sensitivity analysis, in which we excluded all patients with incomplete follow-up for 6MWTD (n=407), which showed consistent findings with our adjusted analysis of the crude population (Supplemental Figure S1). Additional sensitivity analyses excluding all patients with ischemic heart disease (n=164) and atrial fibrillation (n=348) also confirmed our results.

**Figure 3.**
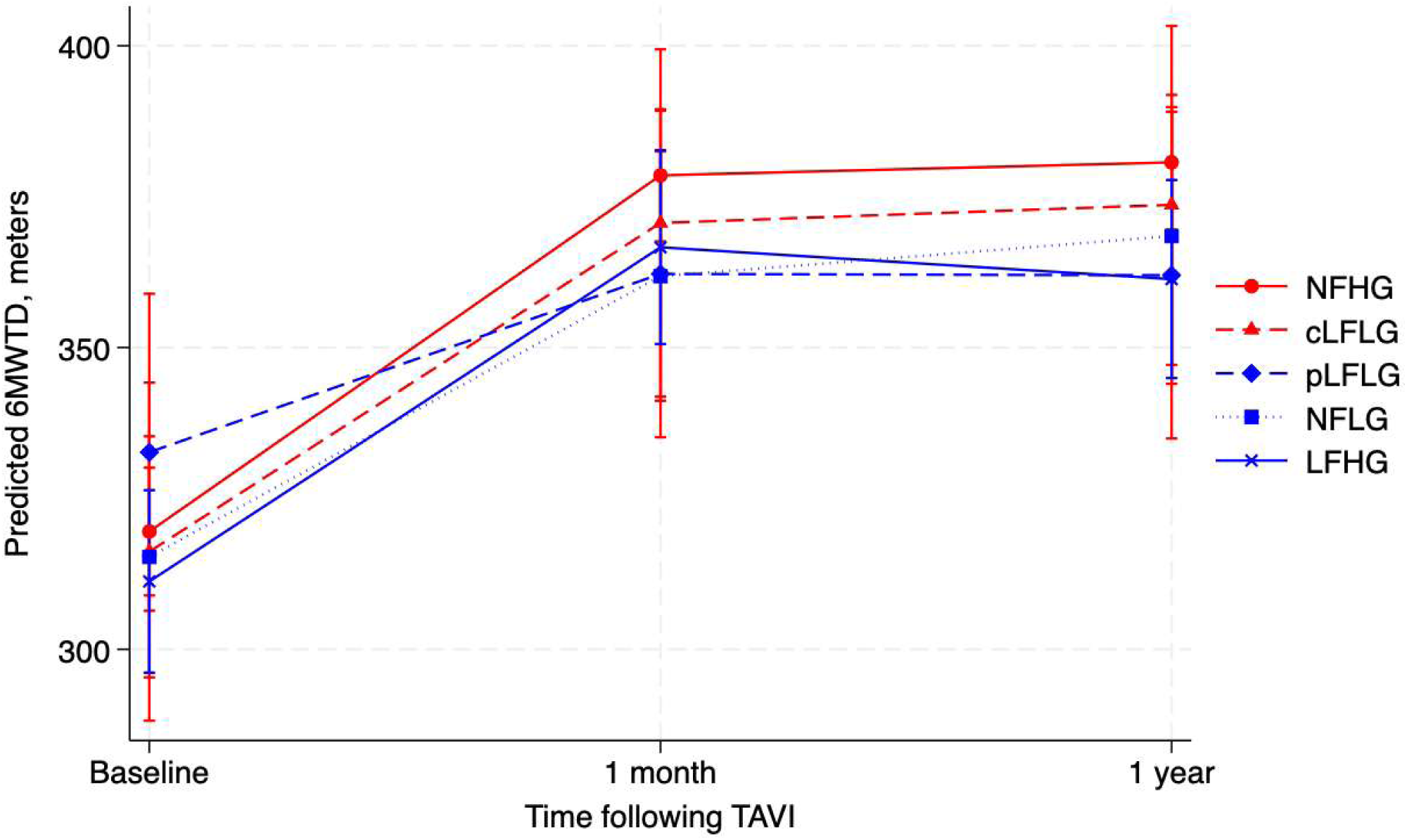
– Adjusted changes in six-minute walk test distance from baseline to 1-month and 1-year follow-up stratified by flow-gradient phenotypes. Covariates for adjustment: Sex, age, BSA, diabetes mellitus, previous MI, AFIB, medical treatment for congestive HF, baseline pacemaker, and NYHA classification. Abbreviations: 6MWTD, six minutes’ walking test distance; AFIB, atrial fibrillation; BSA, body surface area; cLFLG, classical low flow-low gradient; HF, heart failure; LFHG, low flow-high gradient; MI, myocardial infcartion; NFLG, normal flow-low gradient; NFHG, normal flow-high gradient; NYHA, New York Heart Association; pLFLG, paradoxical low flow-low gradient; TAVI, transcatheter aortic valve implantation.

NYHA classification was available for nearly all patients at baseline (99.8%), at 1-month (97.2%), and at 1-year (92.5%) following TAVI. Those with NFLG, cLFLG, and pLFLG AS were more symptomatic at baseline, with NYHA ≥III reported in 40.5%, 57.3%, and 50.6% of patients respectively, compared to 30.5% in patients with normal-flow high-gradient AS (p=0.000, Central illustration). NYHA class improved consistently after TAVI across all groups (p=0.804) (Figure 4), however, 26.2% remained in NYHA class ≥II after 1 year, with significant differences in symptom burden between groups (Central illustration).

**Figure 4.**
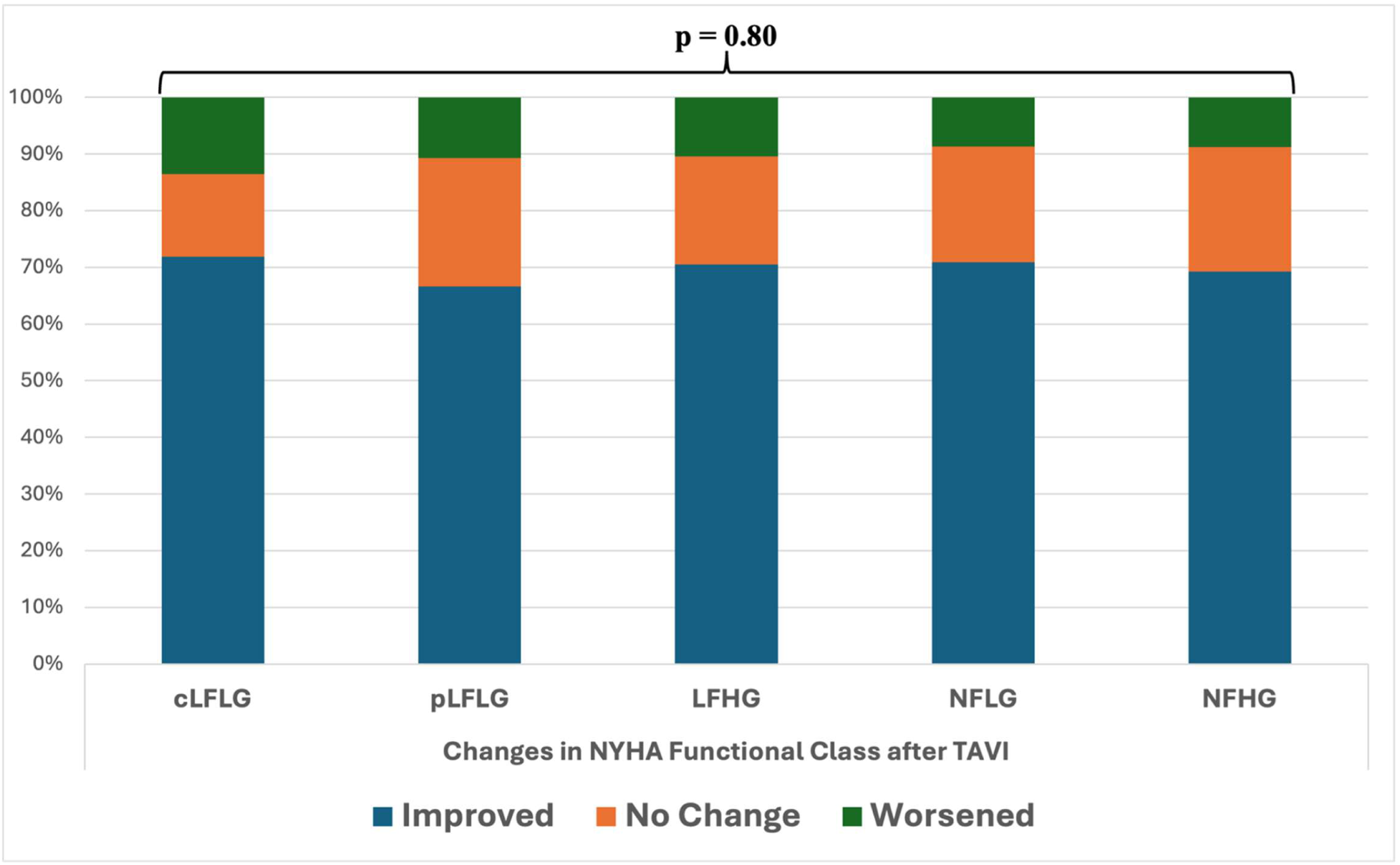
– Histograms illustrating changes in NYHA functional class from baseline until 1 year after TAVI stratified by flow-gradient phenotype. Abbreviations: 6MWTD, six minutes’ walking test distance; cLFLG, classical low flow-low gradient; LFHG, low flow-high gradient; NFLG, normal flow-low gradient; NFHG, normal flow-high gradient; NYHA, New York Heart Association; pLFLG, paradoxical low flow-low gradient; TAVI, transcatheter aortic valve implantation.

NYHA class was already comparable between NFLG and normal-flow high-gradient at 1-month follow-up and persisted after 1 year, whereas patients with cLFLG and pLFLG AS were more likely to have persistent symptoms at 1-month follow-up (NYHA ≥II, 42.7% in both groups, p=0.003). At 1-year follow-up, advanced NYHA class (≥II) was significantly more prevalent in pLFLG (cLFLG 30.3% vs. pLFLG 38.7% vs. low-flow high gradient 27.8% vs. NFLG 29.1% vs. normal-flow high-gradient 21.4%, p=0.012), with no difference between normal-flow high-gradient AS and other flow-gradient phenotypes.

Compared with normal-flow high-gradient AS, patients with NFLG AS demonstrated modest differences in ventricular structure and function at baseline, including larger LV end-diastolic diameter (47.1 mm vs. 45.6 mm, p=0.006) and lower LVEF (56.6% vs. 59.6%, p=0.000) (Figure 6). Patients with cLFLG exhibited the most advanced ventricular remodeling compared to normal-flow high-gradient AS at baseline, with significantly higher LV mass index (133.6 g/m² vs. 120.8 g/m², p=0.000), substantially larger LV end-diastolic diameter (51.9 mm vs. 45.6 mm, p=0.000), and markedly reduced LVEF (35.7% vs. 59.6%, p=0.000). In contrast, patients with pLFLG were characterized by the lowest LV mass index (110.1 g/m², p=0.000), with LV dimensions and LVEF comparable to normal-flow high-gradient AS (Figure 5).

**Figure 5.**
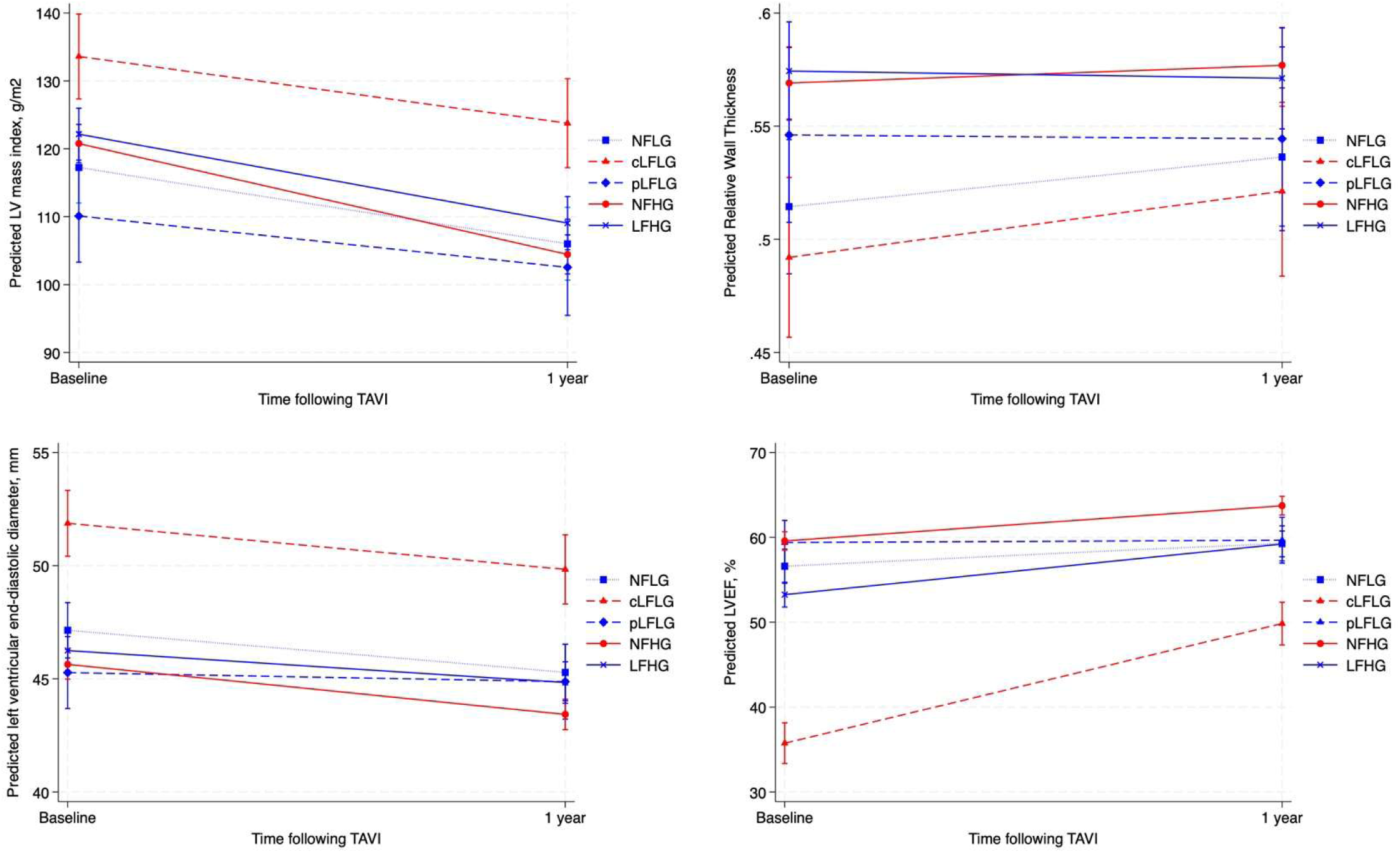
– Changes in reverse remodeling parameters (i.e. LV mass index, LV end-diastolic diameter, relative wall thickness, and LV ejection fraction) from baseline to 1-year follow-up stratified by flow-gradient phenotypes. Abbreviations: cLFLG, classical low flow-low gradient; LFHG, low flow-high gradient; LVEF, left ventricular ejection fraction; NFLG, normal flow-low gradient; NFHG, normal flow-high gradient; pLFLG, paradoxical low flow-low gradient; TAVI, transcatheter aortic valve implantation.

From baseline to 1-year follow-up, LV end-diastolic diameter decreased significantly among patients with normal-flow high-gradient AS (–2.21 mm, p=0.000), with similar reductions observed across all other groups and no evidence of differential changes in LV end-diastolic diameter (Figure 6). LVEF increased significantly in patients with normal-flow high-gradient AS (+4.2%, p=0.000), and comparable increases were observed in patients with NFLG, cLFLG, and low-flow high-gradient AS. In contrast, patients with pLFLG AS demonstrated significantly smaller improvements in LVEF (–3.9%, p=0.029), while relative wall thickness remained unchanged during follow-up across all flow-gradient phenotypes (Figure 5).

After multivariable adjustment (i.e. age, sex, BMI, systolic blood pressure, hypertension, diabetes mellitus, previous myocardial infarction, congestive heart failure, atrial fibrillation, and pacemaker), longitudinal changes in ventricular remodeling parameters remained comparable between normal-flow high-gradient and NFLG AS, while pLFLG AS still demonstrated attenuated LV mass regression, LV end-diastolic diameter reduction, and LVEF improvement.

## DISCUSSION

In this large all-comers study of patients with severe AS undergoing TAVI, we demonstrated that functional recovery and reverse remodeling differ according to flow-gradient patterns following TAVI. Most notably, patients with NFLG demonstrated similar improvements with normal-flow high-gradient AS 1 year after TAVI. These findings are particularly relevant because NFLG AS is a discordant phenotype that is often regarded as moderate AS in contemporary guidelines and is generally not considered an indication for TAVI.^11,14^ In contrast, low-flow low-gradient AS phenotypes demonstrated greater heterogeneity in response to afterload reduction with TAVI; functional recovery among patients with cLFLG AS was comparable to normal-flow high-gradient AS, whereas patients with pLFLG AS demonstrated significantly lower 6MWTD at baseline and during follow-up and remained more symptomatic after 1 year, with approximately 40% reporting persistent symptoms (NYHA class ≥II).

Importantly, the differences in functional recovery were reflected by similar differences in patterns of myocardial reverse remodeling. Patients with normal-flow high-gradient AS demonstrated significant reductions in LV end-diastolic diameter and improvements in LVEF following TAVI, and similar responses to TAVI were observed among patients with NFLG, cLFLG, and low-flow high-gradient AS. Conversely, patients with pLFLG AS exhibited attenuated improvement in LVEF, consistent with limited myocardial reserve. These findings extend the current evidence, suggesting that patients with NFLG have a severe AS phenotype with substantial benefit from afterload reduction with TAVI. In addition, all flow-gradient AS phenotypes shared similar risks of 1-year all-cause death and the composite endpoint of all-cause death, stroke, ≥moderate aortic regurgitation, and ≥moderate hemodynamic valve deterioration.

While it has been accepted for decades that severe AS may occur despite low-gradients in the setting of reduced LVEF,^15^ it was not until the seminal paper by Hachicha and colleagues that it became evident this phenomenon may also occur in patients with preserved LVEF (>50%).^2^ Regardless of LVEF, low-gradient AS raises the diagnostic conundrum of differentiating between severe– and pseudo-severe AS. Currently, guidelines distinguish between patients according to stroke volume.^16^ Patients with low-gradient AS under normal-flow conditions (Svi >35 mL/m^2^) are generally considered to have moderate AS in transition to severe,^9^ whereas further diagnostic testing is required in patients with SV_i_ <35 mL/m^2^ to clarify the severity of AS. In this setting, guidelines recommend the use of an integral approach, often requiring a combination of echocardiographic, clinical and multimodality imaging. If AS is proven severe, aortic valve replacement is recommended.^1^ This recommendation is largely based on retrospective studies reporting a reduced survival along with increased cardiac events among low flow-patients regardless of LVEF.^3,4,9,10,17,18^ Thus, it is intriguing that, despite comparable valvular hemodynamics, our findings demonstrate that patients with cLFLG, and not pLFLG, experience significant functional recovery following TAVI compared to high-gradient AS. While patients with cLFLG may benefit from afterload reduction even when AS is moderate,^19^ this may not be the case in pLFLG where LVEF is preserved.^20^ This interesting difference between low-flow low-gradient groups is probably explained by the fact that patients with reduced LVEF (i.e. cLFLG) have reduced contractility, and thus benefit from afterload reduction,^21,22^ while patients with preserved LVEF (i.e. pLFLG) experience a “heart-failure with preserved ejection fraction”-like physiology that may not improve after TAVI.^5^ We have recently demonstrated that patients with pLFLG were able to increase in stroke volume after TAVI, implying that high filling pressures rather than a reduced cardiac output could be the cause of this lack of improvement (Dyreborg, Mathias, et al, invited for revision following external peer review in JACC: Cardiovascular Imaging). Still our findings are in line with a growing body of evidence demonstrating that low-flow and low-gradients are associated with a futile course after TAVI.^23–26^

Our findings also demonstrate that patients with NFLG exhibit similar improvements in 6MWTD and NYHA functional class to those observed in patients with unequivocally severe AS, defined by the presence of normal-flow high-gradient AS. This finding corroborates previous findings in a study of ours, in which we demonstrated that 87 patients with NFLG referred for surgical aortic valve replacement improved in 6MWTD six month after the procedure, with a subsequent positive myocardial reverse remodeling.^7^ Our findings are also in line with a previous study demonstrating comparable improvements in quality-of-life assessed by questionnaires between patients with NFLG and normal-flow high-gradient AS.^27^ A previous meta-analysis has also demonstrated that patients with NFLG AS referred for aortic valve replacement had better prognosis than patients treated conservatively.^10^ Thus, it is possible that some patients with NFLG may benefit from afterload reduction despite having non-severe AS. Interestingly, it has been shown that an increase in vascular resistance may reduce the mean-gradient, leading to a low-gradient state despite preserved flow and severe AS,^28^ suggesting that this phenotype might benefit from afterload reduction even when AS is not severe. However, it has to be recognized that 50% of patients with NFLG AS have aortic valve calcium scores assessed by CT in the severe range,^29^ and that AS is truly severe with a low-gradient. In our study, patients with NFLG AS also had a mean AVA of 0.93 cm^2^ (SD 0.18), placing them in the severe rather than moderate AS category. Future studies should be conducted to assess the impact of aortic valve replacement in these patients.

### Limitations

Several limitations warrant consideration. First and most important we did not have a comparison group of patients undergoing sham procedure. This may imply that functional recovery may partially reflect a placebo-related effect. It is thus important that we also demonstrate a difference in 6MWTD and reverse remodeling parameters, as these objective changes are less likely to be placebo related.

Second, we did not routinely test for cardiac amyloidosis, and thus, we cannot rule out that a part of the lack of improvement seen in pLFLG patients reflects undiagnosed concomitant cardiac amyloidosis.

Third, data from the COMPARE-TAVI 1 trial represent a multi-center, single-country setting, which may limit the external validity of our findings to other countries. However, tertiary centers follow the guidelines of the Danish Society of Cardiology, which endorse the international guidelines for AS published by the European Society of Cardiology and American College of Cardiology.

Fourth, nearly 50% of data for 6MWTD was missing at 1-year follow-up. Given that missing data is likely associated with adverse outcomes such as death or significant functional decline, the data may not be missing at random. This introduces the potential for bias, as patients with the worst functional trajectories may be underrepresented in each flow-gradient phenotype. However, multivariable adjustments for clinically relevant parameters and sensitivity analyses excluding all patients with incomplete follow-up for 6MWTD confirmed robustness of our results, supporting the overall findings in our sub-study.

## CONCLUSION

TAVI improves functional capacity measured as changes in NYHA functional class and 6MWTD in all flow-gradient phenotypes, although with significant heterogeneity between groups. While patients with NFLG shared the improvements in functional capacity and reverse remodeling of those with high-gradient AS, patients with pLFLG remained more symptomatic with attenuated improvements in 6MWTD and reverse remodeling at 1-year follow-up.

## Data Availability

All study-related documents will be made available on request. Individual data collected for the study will be made available for collaborative pooled analyses provided relevant contracts and data sharing agreements are made. Only anonymized data will be shared. Any requests for data access should be directed to the sponsor at Aarhus University via.

## ACKNOWLEDGEMENTS

Data management was provided and REDCap was hosted by OPEN, Open Patient data Explorative Network, Odense University Hospital, Region of Southern Denmark. For the main study (COMPARE-TAVI 1) data was hosted at Aarhus University Hospital in corolog.net. During the preparation of this work the author(s) occasionally utilized ChatGPT for grammar assistance. After using this tool/service, the author(s) reviewed and edited the content as needed and take(s) full responsibility for the content of the publication.

## ROLES AND SOURCES OF FUNDING

The COMPARE-TAVI 1 trial was funded by Meril Life Sciences, Vingmed A/S Denmark, the Danish Heart Foundation, and the Central Denmark Region. The funding sources had no influence on the protocol, conduct of this sub-study, or submission of the results for publication. The funding sources had no access to trial data and were not informed about results of the study before submission of the results.^30^

## ABBREVIATIONS

AS: Aortic stenosis
LV: Left ventricle
AVA: Aortic valve area
SVi: Stroke volume index
LVEF: Left ventricular ejection fraction
TAVI: Transcatheter aortic valve implantation
NFLG: Normal flow-low gradient
NYHA: New York Heart Association
6MWTD: Six-minutes’ walk test distance
cLFLG: Classical low flow-low gradient
pLFLG: Paradoxical low flow-low gradient

## Figure legends

**Central illustration.**
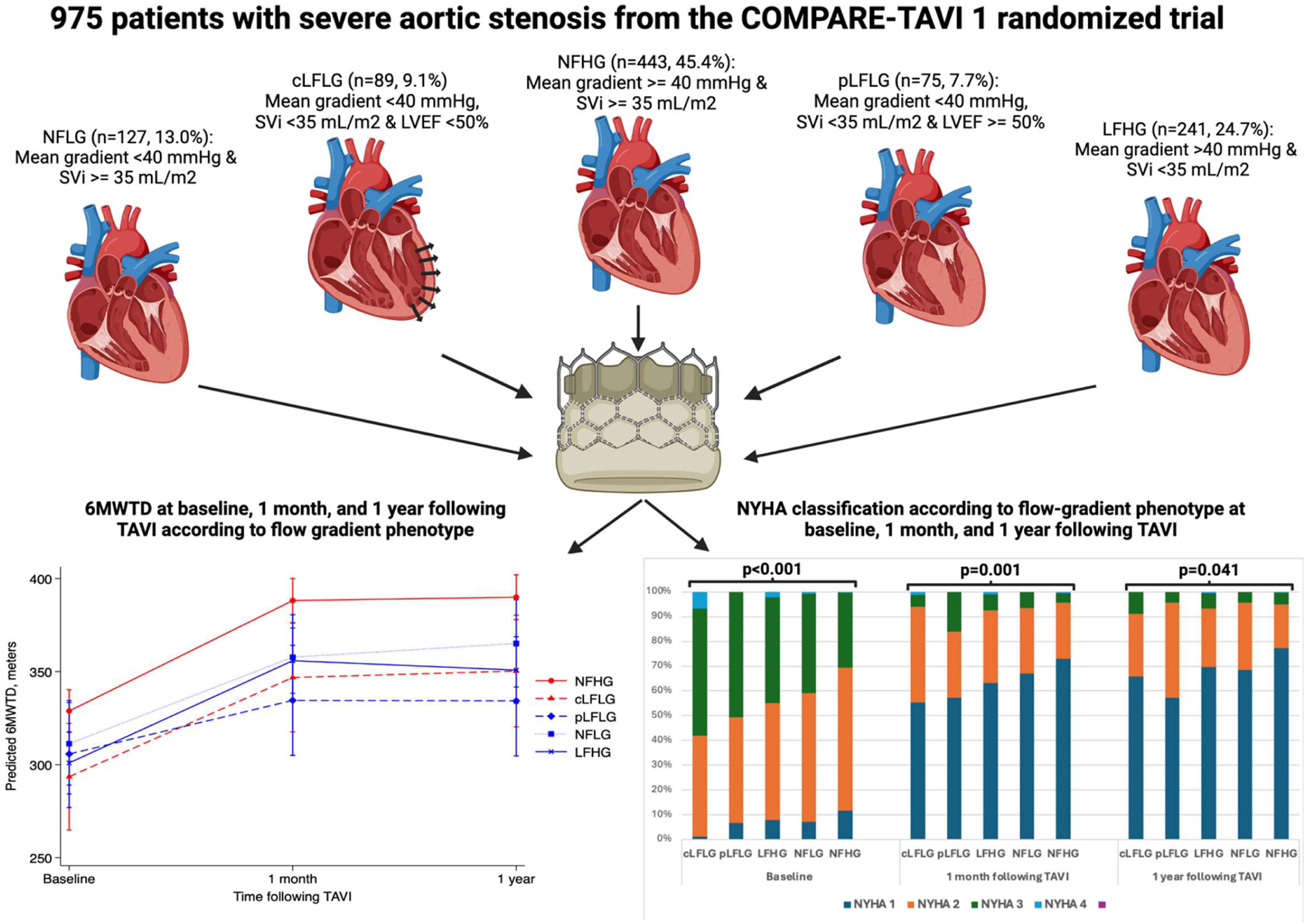
– Patients with severe aortic stenosis stratified in subgroups according to aortic valve area, stroke volume, mean gradient, and left ventricular ejection fraction. Improvement in six-minute walk test distance following TAVI demonstrated some heterogeneity with the comparable improvements between normal-flow high-gradient AS and NFLG AS, and the least increase was seen in patients with pLFLG AS. In addition, patients in this group were the most symptomatic 1 year following TAVI. This illustration was created with Biorender.com Abbreviations: 6MWTD, six minutes’ walk test distance; cLFLG, classical low flow-low gradient; LFHG, low flow-high gradient; NFLG, normal flow-low gradient; NFHG, normal flow-high gradient; NYHA, New York Heart Association; pLFLG, paradoxical low flow-low gradient; TAVI, transcatheter aortic valve implantation.

